# A comparison of clinical outcomes, service satisfaction, and well-being in people using Acute Day Units and Crisis Resolution Teams: a cohort study in England

**DOI:** 10.1101/2020.11.17.20233254

**Authors:** Danielle Lamb, Thomas Steare, Louise Marston, Alastair Canaway, Sonia Johnson, James B. Kirkbride, Brynmor Lloyd-Evans, Nicola Morant, Vanessa Pinfold, Deb Smith, Scott Weich, David Osborn

## Abstract

**Background:** For people in mental health crisis, Acute Day Units (ADUs) provide daily structured sessions and peer support in non-residential settings, often as an addition or alternative to Crisis Resolution Teams (CRTs). There is little recent evidence about outcomes for those using ADUs, particularly in comparison to those receiving CRT care alone.

**Aims:** To investigate readmission rates, satisfaction, and wellbeing outcomes for ADU and CRT service users.

**Methods:** A cohort study comparing readmission to acute mental health care during a six-month period for ADU and CRT participants. Secondary outcomes included satisfaction (CSQ), wellbeing (SWEMWBS), and depression (CES-D).

**Results:** We recruited 744 participants (ADU: 431, 58%; CRT 312, 42%) across 4 NHS Trusts/health regions. There was no statistically significant overall difference in readmissions; 21% of ADU participants (versus 23% CRT) were readmitted over 6 months (adjusted HR 0.78, 95%CI 0.54, 1.14). However, readmission results varied substantially by setting. At follow-up, ADU participants had significantly higher Client Satisfaction Questionnaire (CSQ) scores (2.5, 95% CI 1.4 to 3.5, p<0.001) and wellbeing scores (1.3, 95%CI 0.4 to 2.1, p=0.004), and lower depression scores (−1.7, 95%CI −2.7 to −0.8, p<0.001) than CRT participants.

**Conclusions:** Service users who accessed ADUs demonstrated better outcomes for satisfaction, wellbeing, and depression, and no significant differences in risk of readmission compared to those who only used CRTs. Given the positive outcomes for service users, and the fact that ADUs are inconsistently provided across the country, their value and place in the acute care pathway needs further consideration and research.

## Introduction

Within the NHS, support for people in mental health crisis is typically provided by multidisciplinary Crisis Resolution Teams (CRTs), which aim to avoid inpatient admission by providing care at home via frequent visits. However, there is evidence that implementation of national guidelines for CRTs is highly variable, meaning that some people may not receive the intensity of support they need,^1^ with lack of therapeutic content and social contact frequently raised as issues.^2^ In addition, CRT care is often dependent on support from carers, which is problematic where there are no carers, and can lead to excessive burden even where there are carers.^3^

Acute Day Units (ADUs), previously known as ‘day hospitals’,^4^ provide an additional clinical resource for those in mental health crisis. In England, there is no NHS-specified model, but ADUs typically offer on-site individual and group sessions during the day, with service users returning home overnight and at weekends. ADU care is provided by a multidisciplinary team, usually including nurses, therapists, psychiatrists and other mental health professionals^5^. As a result, service users are provided with structured days, more staff contact time and continuity than is available via CRTs, with opportunities for peer support from other clients, and a wider range of psychological, social and medical interventions. ADUs are often used concurrently with other services (e.g. CRTs, inpatient wards, crisis houses), with these services all referring to each other. Evidence of the effectiveness of ADUs is limited, with the most recent meta-analysis conducted in 2011.^4^ This review synthesised evidence from studies that compared ADUs with hospital admission and concluded that they provide a viable alternative to hospital admission for some, with similar effectiveness on readmission rates after discharge, employment, quality of life, and treatment satisfaction, but quality of evidence overall was reported as low.^4^ Furthermore, to date there has been no comparison of outcomes and experiences of ADU service users compared with those using CRTs, arguably a more directly comparable type of service.

We need evidence regarding any additional benefit ADUs may offer to service users, not only in terms of clinical outcomes, but also on patient-reported outcomes such as patient experience, wellbeing, and quality of life. The aims of this study were:

1. To describe and compare the clinical and socio-demographic characteristics of people using ADUs and CRTs.
2. To compare outcomes in terms of readmission, wellbeing, depression, and service user satisfaction for those who received ADU care with those who received only CRT care.

We hypothesised that people receiving ADU care would have fewer admissions, greater satisfaction and wellbeing, and less depression at 6 months, compared with those receiving CRT care alone.

## Methods – cohort study

The authors assert that all procedures contributing to this work comply with the ethical standards of the relevant national and institutional committees on human experimentation and with the Helsinki Declaration of 1975, as revised in 2008. All procedures were approved by the London Bloomsbury Research Ethics Committee (ref: 16/LO/2160).

### Design and setting

We established a cohort study of ADU and CRT users and compared readmission to the acute care pathway during a six-month period from baseline, as well as measures of depression, wellbeing, and satisfaction with services. We recruited four NHS sites with ADUs and CRTs in England. Participant recruitment took place between March 2017 and April 2019, with follow up completed in September 2019.

We invited people consecutively admitted to each service to participate in baseline interviews. Recruitment and data collection could occur at any point during the initial admission, and up to 14 days post-discharge from the service. Telephone or online follow-up was carried out 8-12 weeks post-baseline, with electronic health records outcome data collected 6 months post-baseline.

## Participants

### Inclusion and exclusion criteria

Inclusion criteria were as follows:

- 18 years old or older
- Used an ADU/CRT service for at least one week
- Read and understand English (or translator available)
- Capacity to provide informed consent
- Did not pose too high a risk to others or themselves to participate (as judged by their current clinical team)

ADU participants could use CRTs concurrently or during the follow up period, however CRT participants were excluded if they used an ADU at any point during study period. This was to determine benefits of ADUs as an addition to the acute care pathway, over and above CRTs.

## Measures

### Exposure

Our main exposure was being under the care of an ADU (solely or in combination with CRT use) for at least one week before baseline, versus CRT care only.

### Outcomes

Our primary outcome was time to readmission for acute treatment, after discharge from CRT or ADU, during the 6-month study period. This was collected via service use data from Electronic Health Records (EHRs). We defined readmission for acute treatment as any subsequent use of acute mental health services (CRT, crisis house, ADU, or inpatient ward) after discharge from the service used at baseline during the subsequent 6-month study period.

Our secondary outcomes were self-reported satisfaction with mental health services (Client Satisfaction Questionnaire; CSQ),^6^ wellbeing (Short Warwick Edinburgh Mental Well Being Scale; SWEMWBS),^7^ and depression (Center for Epidemiologic Studies Depression Scale; CES-D),^8^ collected via online questionnaire at baseline and 8-12 weeks later.

### Covariates

We collected demographic data and self-rated physical health via the baseline questionnaire. We collected data from EHRs on: admissions and service use; clinical characteristics (ICD10 diagnosis and any comorbid diagnoses, physical health diagnoses, substance misuse, smoking, medication, previous inpatient use); Health of the Nation Outcome Scale (HoNOS) scores;^9^ and content of care (physical assessment carried out, carers involved in care, psychological input from service used at baseline). Where service users had multiple diagnoses recorded in their EHR, we recorded the diagnosis considered to be more severe. We recorded whether the person had a Serious Mental Illness (SMI), typically defined as being diagnosed with Schizophrenia, other non-organic Psychoses, or Bipolar Disorder.^10^

### Procedure

At baseline, ADU/CRT staff screened all service users consecutively admitted to their service from the study start date. All service users who met the inclusion criteria were approached by clinical or research staff and asked if they were willing to discuss participation further (except at sites where service users had already given consent to be contacted directly about research projects: in this instance, researchers contacted service users directly once their eligibility and any risk-related safety requirements had been established from clinicians and patient records). Those who agreed to discuss the study were contacted by a researcher with an information sheet and an offer to answer any questions. Potential participants were given at least 24 hours to consider whether they would like to take part, and then if still interested they provided written consent to a researcher, who also collected the baseline data. Consent and data collection could occur up to 14 days’ post-discharge from initial service use.

Participants were offered £20 (vouchers) reimbursement for taking part (£10 for the baseline interview, and £10 for the follow up interview at 8-12 weeks after baseline). Participants were contacted by phone by a researcher 8-12 weeks after baseline to collect follow up data. At 6 months after baseline, readmission data were collected from EHRs.

### Sample size

*A priori*, we estimated that a sample size of 400 patients per group (N=800) would give 90% power to detect a difference of 12% in the proportions readmitted in each arm (with an assumption of 50% readmission in the CRT group), with alpha set to 0.05. It would also afford 90% power to detect an effect size difference of 0.3 on the CSQ. This calculation included inflation for clustering by Trust.

### Analysis

We calculated descriptive statistics comparing the baseline characteristics of ADU versus CRT users for the sample as a whole, and within sites. We explored baseline differences in demographics, clinical data, and content of care, using parametric and non-parametric tests as appropriate, as well as the proportion of ADU versus CRT users admitted during the 6-month study period.

For our primary outcome we compared time to readmission in ADU and CRT participants. We analysed this using Cox’s regression to produce a hazard ratio. Cohort entry was the date of recruitment and cohort exit date was the date of readmission to acute care or the 6-month study end point. We adjusted for variables chosen *a priori*, which previous research suggested may be relevant (Trust, age, sex, SMI diagnosis, employment, baseline HoNOS, baseline SWEMWBS, and whether or not the person had previously been an inpatient). Covariates were added using a stepwise procedure, starting with a bivariate model with Trust as a fixed effect, then adding the variables of age, sex, and employment, followed by diagnosis, followed by HoNOS, followed by SWEMWBS, followed by previous inpatient. Due to expected heterogeneity between different Trusts, we tested for an interaction between type of team (ADU/CRT) and Trust as a sensitivity analysis.

For the secondary outcomes we analysed mean satisfaction, wellbeing, and depression scores at weeks 8-12 using linear regression. Stata 16 was used for all analyses.^11^

## Results – cohort study

Figure 1 below shows the flow of participants into the study, and those followed up at 8-12 weeks. Only one participant declined consent for access to their EHR at the 6-month follow up, meaning a completion rate of >99% for the primary outcome of readmission.

**Figure 1.**
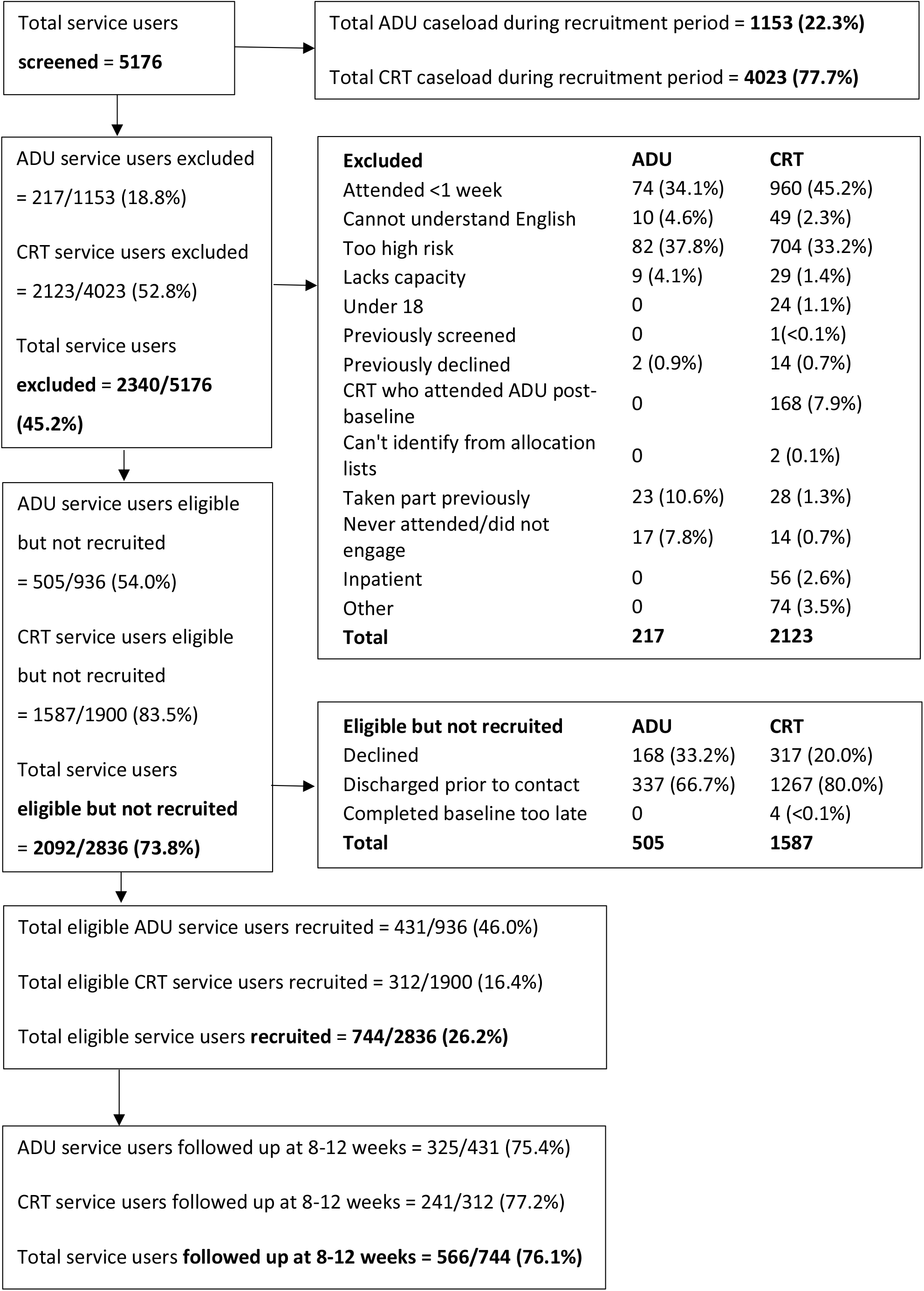
STROBE diagram of the flow of participants into the study

### Cohort descriptives – baseline sociodemographic and clinical characteristics

We recruited 744 participants, of whom 431 (57.9%) had received ADU care and 312 (42.1%) had only received CRT care. Due to small numbers of black and ethnic minority participants we have used undesirably broad ethnic categories. The ADU and CRT groups were generally similar in terms of sociodemographic characteristics (full details available in Table 1). Those sociodemographic characteristics with statistically significant differences between the groups are outlined below.

**Table 1.** Sociodemographic characteristics, clinical characteristics, and baseline measures

|  | ADU | CRT | Significance |
| --- | --- | --- | --- |
| **Sociodemographics** |  |  |  |
| Age<br>mean(SD) | 41.8 (14.0) | 39.2 (13.0) | Mean diff = -2.6 |
|  |  |  | 95%CI -4.6 to -0.5<br>p=0.01 |
| Sex |  |  |  |
| Male | 220 (51.0) | 140 (44.9) | p=0.1 |
| Female | 211 (49.0) | 172 (55.1) |  |
| Ethnicity |  |  |  |
| White (UK & non-UK) | 362 (84.6) | 251 (82.3) | p=0.2 |
| Black (UK, African, Caribbean, Other) | 23 (5.4) | 17 (5.6) |  |
| Asian (UK, south Asian, Chinese, Other) | 28 (6.5) | 17 (5.6) |  |
| Mixed | 15 (3.5) | 20 (6.6) |  |
| Employed (Yes) | 126 (29.2) | 133 (42.6) | p<0.001 |
| Marital status |  |  |  |
| Single | 268 (62.2) | 189 (60.6) | p=0.4 |
| Cohabiting | 41 (9.5) | 37 (11.9) |  |
| Married | 88 (20.4) | 69 (22.1) |  |
| Divorced | 31 (7.2) | 15 (4.8) |  |
| Widowed | 3 (0.7) | 2 (0.6) |  |
| Clinical Characteristics |  |  |  |
| ICD10 primary diagnosis |  |  |  |
| Psychosis | 80 (19.1) | 40 (14.0) | p=0.01 |
| Mood disorders | 210 (50.0) | 143 (50.2) |  |
| Anxiety | 55 (13.1) | 55 (19.3) |  |
| Personality disorders | 63 (15.0) | 30 (10.5) |  |
| Other | 12 (2.9) | 17 (6.0) |  |
| SMI (Yes) | 139 (33.1) | 80 (28.1) | p=0.2 |
| Physical health |  |  |  |
| Excellent | 21 (4.9) | 17 (5.5) | p=0.2 |
| Very good | 84 (19.5) | 49 (15.7) |  |
| Good | 147 (34.1) | 101 (32.4) |  |
| Fair | 124 (28.8) | 85 (27.2) |  |
| Poor | 55 (12.8) | 60 (19.2) |  |
| Comorbidity |  |  |  |
| Mental health | 100 (23.6) | 74 (23.9) | p=0.1 |
| Physical health | 101 (23.8) | 57 (18.4) |  |
| Both | 58 (13.7) | 38 (12.3) |  |
| Substance misuse (Yes) | 103 (24.3) | 85 (27.6) | p=0.5 |
| Smoker (Yes) | 172 (41.1) | 114 (38.4) | p=0.4 |
| Previous inpatient (Yes) | 248 (57.5) | 116 (37.2) | p<0.001 |
| # previous admissions (mean/SD) | 1.44 (2.5) | 1.09 (3.0) | Mean diff = -0.3<br>95%CI -0.8 to 0.1 |
| **Content of care** |  |  |  |
| Physical assessment | 340 (80.4) | 106 (34.2) | p<0.001 |
| Carers involved | 183 (43.1) | 102 (32.9) | p=0.01 |
| Psychological input | 248 (58.8) | 174 (41.2) | p<0.001 |
| **Baseline measures** |  |  |  |
| CES Depression (mean/SD) | 16.4 (5.3) | 17.5 (5.4) | Mean diff = 1.1<br>95%CI 0.4 to 2.0<br>p=0.009 |
| SWEMWBS (mean/SD) | 20.5 (5.0) | 19.3 (5.0) | Mean diff = -1.2<br>95%CI -2.1 to -0.4<br>p=0.004 |
| HoNOS (mean/SD) | 14.1 (6.1) | 12.43 (6.2) | Mean diff = -1.7<br>95%CI -2.6 to -0.7<br>p<0.001 |

ADU and CRT data are n (%) unless otherwise specified.

ADU participants were older than CRT participants, and less likely to be employed. Clinically, higher proportions of ADU participants were diagnosed with psychosis and personality disorders, and a higher proportion of CRT participants were diagnosed with anxiety disorders. A higher proportion of ADU participants had previously been admitted to a psychiatric inpatient ward.

In terms of content of care while using the ADU or CRT, a much larger proportion of ADU participants received a physical health assessment, had carers involved in their care, and received psychological input, compared to CRT participants.

At baseline CES Depression scores were lower in ADU participants than in CRT, while SWEMWBS wellbeing scores were higher in ADU than CRT participants. The mean baseline total HoNOS score was higher for ADU participants than for CRT participants, indicating more severe difficulties for ADU participants.

There were slight differences in the length of index admission and time at which participants were recruited. ADU participants had a mean index admission lasting for 60 days (SD 38) before they were discharged, while CRT participants had a mean index admission of 43 days (SD 39) before discharge. The mean difference in length of index admission between ADU and CRT participants was-17 days, which was statistically significant (p<0.0001, 95%CI −22.0 to −11.1). ADU participants were recruited at a mean of 24 days (SD 23) into their index admission (i.e. 24 days after being admitted to the service they were recruited from), and CRT participants were recruited a mean of 31 days (SD 24) into their index admission. The mean difference between ADU and CRT participants in time from index admission to recruited was 7 days, which was statistically significant (p=0.0001, 95%CI 3.2 to 9.8).

Follow-up data were collected for >99% of participants for the primary outcome, and 76% of participants for the secondary outcomes. The only significant difference between those who did and did not complete the secondary outcome questionnaires was that completers were older (mean age 41.6, SD 13.7) than those lost to follow-up (median age 37.8, SD 13.0) (p=0.001, 95%CI −6.2 to −1.5).

### Primary outcome – readmission to acute care over 6-month follow-up

For the primary outcome at the 6-month time point, 21.4% of ADU participants were readmitted, compared with 23.4% of CRT participants, with no statistically significant difference evident. The results from the Cox regression comparing time to admission in ADU versus CRT participants are shown in Table 2. Model 1 shows a bivariate analysis, including only team and Trust. Model 2 adjusted for age, sex, and employment, while Model 3 added the remaining covariates (SMI diagnosis; whether participants had previously been an inpatient; baseline HoNOS score; baseline SWEMWBS score).

**Table 2.** Primary outcome – hazard ratios of readmission to acute care comparing ADU and CRT participants

|  | **Model 1 –**<br>**team, Trust** | **Model 2 – age,**<br>**sex,**<br>**employment** | **Model 3 – SMI,**<br>**previous**<br>**inpatient,**<br>**HoNOS,**<br>**SWEMWBS** | **Model 5 –**<br>**interaction**<br>**between team**<br>**and Trust** |
| --- | --- | --- | --- | --- |
| CRT | 1 | 1 | 1 | - |
| ADU | 0.94 (0.67, 1.32) | 0.90 (0.64, 1.28) | 0.78 (0.54, 1.14) | - |
| Trust 1 | 1 | 1 | 1 | 1 |
| Trust 2 | 0.54 (0.37, 0.81) | 0.52 (0.35, 0.77) | 0.52 (0.34, 0.79) | 0.41 (0.22, 0.75) |
| Trust 3 | 0.31 (0.19, 0.49) | 0.31 (0.19, 0.49) | 0.46 (0.27, 0.77) | 0.04 (0.01, 0.32) |
| Trust 4 | 0.15 (0.83, 0.28) | 0.15 (0.08, 0.28) | 0.17 (0.09, 0.32) | 0.13 (0.05, 0.34) |
| Age | - | 0.99 (0.98, 1.01) | 0.99 (0.98, 1.01) | 0.99 (0.98, 1.01) |
| Male | - | 1 | 1 | 1 |
| Female | - | 1.18 (0.84, 1.66) | 1.41 (0.98, 2.04) | 1.42 (0.99, 2.04) |
| Not employed | - | 1 | 1 | 1 |
| Employed | - | 0.78 (0.53, 1.13) | 0.92 (0.61, 1.39) | 0.88 (0.58, 1.33) |
| No SMI | - | - | 1 | 1 |
| SMI | - | - | 1.51 (1.03, 2.23) | 1.45 (0.98, 2.13) |
| Not previous inpatient | - | - | 1 | 1 |
| Previous inpatient | - | - | 2.65 (1.72, 4.07) | 2.31 (1.51, 3.52) |
| HoNOS | - | - | 1.03 (1.00, 1.06) | 1.02 (0.99, 1.05) |
| SWEMWBS | - | - | 0.99 (0.94, 1.03) | 0.99 (0.94, 1.04) |
| Team#Trust1 | - | - | - | 0.46 (0.25, 0.84) |
| Team#Trust2 | - | - | - | 0.74 (0.39, 1.43) |
| Team#Trust3 | - | - | - | 9.11 (1.22, 68.09) |
| Team#Trust4 | - | - | - | 0.83 (0.27, 2.54) |

Data are hazard ratio (95% confidence interval), fully adjusted for all covariates.

We included a cross-site interaction between team (ADU or CRT) and Trust in Model 4, which showed there were significant differences across sites in terms of the primary outcome of readmission (Likelihood Ratio Test p<0.001). Model 4 showed that within Trust 1 the risk of readmission was statistically significantly lower for ADU participants than CRT participants. In Trusts 2 and 4 there was no statistically significantly difference in risk of readmission. In Trust 3 the risk of admission was statistically significantly higher for ADU participants than CRT participants. Full results are given in Table 2.

### Secondary outcomes – satisfaction, wellbeing, and depression at 8-12 weeks

We found statistically significantly higher service user satisfaction (CSQ) scores and wellbeing scores (SWEMWBS), and lower levels of depression (CES-D) in ADU participants than CRT participants at the 8-12 week follow-up period. The full results the linear regressions examining these differences in fully adjusted models are shown in Table 3.

**Table 3.** Secondary outcomes – results from linear regression of satisfaction, wellbeing, and depression at 8-12 weeks

|  | **CSQ**<br>Scoring scale 8-32 | **SWEMWBS**<br>Scoring scale 7-35 | **CES-D**<br>Scoring scale 0-30 |
| --- | --- | --- | --- |
| Team (CRT) |  |  |  |
| ADU | 2.5 (1.4 to 3.5) | 1.3 (0.4 to 2.1) | -1.7 (-2.7 to -0.8) |
| Trust (Trust 1) |  |  |  |
| Trust 2 | -1.1 (-2.7 to 0.6) | -1.4 (-2.7 to -0.1) | 1.2 (-0.2 to 2.6) |
| Trust 3 | -0.5 (-2.2 to 1.2) | -0.1 (-1.4 to 1.2) | 1.7 (0.2 to 3.1) |
| Trust 4 | -0.1 (-1.7 to 1.5) | -0.3 (-1.6 to 1.0) | -1.0 (-2.4 to 0.4) |
| Age | 0.1 (0.0 to 0.1) | 0.02 (-0.01 to 0.05) | 0.007 (-0.04 to 0.03) |
| Sex (Male) |  |  |  |
| Female | -0.1 (-1.1 to 0.9) | 0.02 (-0.01 to 0.1) | 0.6 (-0.3 to 1.5) |
| Employment (None) |  |  |  |
| Employed | 1.9 (0.8 to 3.0) | 1.8 (1.0 to 2.7) | -2.0 (-3.1 to -1.1) |
| SMI (None) |  |  |  |
| SMI | 0.1 (-1.1 to 1.3) | 0.004 (-1.0 to 1.0) | -1.3 (-2.4 to -0.3) |
| Previous inpatient (None) |  |  |  |
| Previous inpatient | 0.01 (-1.0 to 1.2) | -0.2 (-0.7 to 1.0) | 0.03 (-1.0 to 0.9) |
| HoNOS | -0.1 (-0.2 to 0.0) | -0.1 (-0.1 to 0.0) | 0.05 (0.03 to 0.1) |
| SWEMWBS baseline | - | 0.5 (0.4 to 0.6) | - |
| CES-D baseline | - | - | 0.5 (0.4 to 0.6) |

Data are coefficients from linear regression (95% confidence interval), fully adjusted for all covariates.
Interpretation: CSQ and SWEMWBS are positively scored (higher score = better), CES-D is negatively scored (lower = better).

## Discussion

### Summary of results

In a fully adjusted model there was no significant difference between ADU and CRT participants in terms of readmission. However, once an interaction effect was included in the analysis, significant differences between ADU and CRT participants became apparent, with ADU participants in different Trusts being variously at increased, decreased, or no different risk of readmission compared to CRT participants. At 8-12 weeks, after accounting for baseline differences in participant characteristics, ADU participants had significantly higher satisfaction scores, with better wellbeing and lower depression scores than CRT participants. These results indicate that, despite serving a more unwell client group (as suggested by the lower proportion of ADU participants in employment, and higher proportions with SMI and previous inpatient use), overall, ADUs produce comparable outcomes in terms of readmission, and better satisfaction, depression outcomes, and wellbeing than CRTs.

### Comparison to previous research

In terms of the primary outcome, readmission to acute services, we are not aware of previous research directly comparing ADU and CRT services. The most recent meta-analysis of research on ADUs^4^ found no difference in subsequent readmission rates between people using ADU and inpatient services. However, this meta-analysis did find some evidence (albeit low quality) that those using ADUs had better outcomes than inpatients regarding subsequent employment. This is particularly interesting given our finding that ADU participants in this cohort had lower rates of employment than CRT participants. One study investigated readmission of CRT service users over 1 year,^12^ and found that psychotic disorders increased the risk of readmission. Their study found similar HoNOS scores in two CRT cohorts to our CRT participants (13.3, SD 6.4, and 11.8 SD 5.5). The sample used in that study was similar in terms of sex (52% female) though with a smaller number of white participants (68%). Similarly, another study of CRT service users found a readmission rate of 38%,^13^ which is comparable to the current study. That study found similar CSQ scores in CRT service users (mean score of 26, SD 5) to the current study, although the sample had a slightly different demographic profile (female 60% vs 51% in the current study; white ethnicity 65% vs 83% in the current study).^13^

We are not aware of previous research of ADU and CRT service users directly comparing the two groups on the secondary outcomes we used. Previous studies have compared outcomes for ADUs and inpatient wards, and satisfaction and readmissions among patients treated by CRTs. In a randomised controlled trial comparing day hospitals to inpatient care,^14^ satisfaction was measured and, similar to this study, found significantly higher satisfaction in the day hospital group than the inpatients group at discharge and three months (though no difference at 12 months). The CSQ-8 has been used in several other studies of acute mental health services, for example, in a cohort study of people using standard acute inpatient wards compared to those using alternative residential crisis services (e.g. crisis houses), those using alternative services had significantly higher satisfaction scores (26.4, SD 4.9, versus 23.1, SD 6.6).^15^ These scores are comparable with those found in the current study (26.66, SD 5.04, versus 24.37, SD 6.57), with greater satisfaction among those using ADUs and crisis houses than those using CRTs and inpatient wards.

### Strengths and limitations

There are four key strengths of this cohort study: 1) the direct comparison of those using ADUs and CRTs, which has not been undertaken previously; 2) the robust methodology (large sample size and adjustment for clinical and demographic differences at baseline); 3) the range of geographical locations of participating services; 4) high follow-up rates. As discussed above, there is a lack of evidence about ADUs in modern mental health settings, with the majority of work having been carried out some time ago, and on services that were substantially different to those available currently. This study provides a comprehensive overview of the sociodemographic and clinical characteristics of those using ADUs and CRTs, and offers insights to the comparative benefits of each type of service in terms of clinical and wellbeing outcomes. To our knowledge, this is the largest cohort study of ADU users to date, and methodologically rigorous, lending weight to these results.

The participating services are located in areas serving urban, suburban, and rural communities, with high and low deprivation levels. We were able to follow up 99.99% of participants via their EHRs at the 6-month time point, which included our primary outcome of service use during the 6 months, and 76% of participants (ADU 75.4%; CRT 77.2%) at the 8-12 week time point, which included the secondary outcomes of satisfaction, wellbeing, and depression.

There are four main limitations of the study: 1) selection and attrition bias; 2) differences in index admission and recruitment periods; 3) power and lack of randomisation of participants; 4) EHR data quality. Firstly, as is typical in cohort studies of this nature, we were reliant on people who were attending participating services being willing and able to take part. The nature and severity of illness of some service users excluded them from the study – around a third of potential participants were excluded (from both service types) due to risk considerations – and as a result our findings may not be applicable to the most unwell users of these services (which is a potential issue for any type of study). However, the sample recruited from both ADUs and CRTs included a variety of diagnoses including people who had previously been admitted to inpatient care, and both groups had substantial use of mental health services at follow-up. Furthermore, many participants were single, unemployed, and diagnoses included psychosis and personality disorder. This indicates that our sample were indeed people with complex mental health needs. That said, the overall recruitment rate from those eligible for inclusion was 46% in ADUs and only 16% in CRTs. In part this likely reflects the fact that face to face recruitment (possible in ADUs but not CRTs) tends to be easier than recruitment over the phone (the only recruitment method used in CRTs). As a result, it may be that the CRT group in this sample is unrepresentative. However, the relatively high response rate at follow up (76%), and lack of significant differences between those completing the follow up questionnaire and those not completing, can give us confidence that attrition bias is not a concern.

Secondly, there were statistically significant differences between ADU and CRT participants in terms of the lengths of their index admission (with ADU participants on average being admitted for 17 days longer than CRT participants), and in terms of the time from index admission to recruitment (with CRT participants recruited an average of 7 days later in their index admission than ADU participants). The longer length of admission in ADUs could reflect the fact that this group appears to be more severely unwell, with higher rates of SMI and previous inpatient admission, and lower rates of employment. Although there was a significant difference in time from index admission to recruitment, this was relatively small (7 days), and seems unlikely to have had a substantial impact on our primary or secondary outcomes.

Thirdly, we recruited 744 people to the study, having originally aimed for a target of 800, which would have given 90% statistical power to detect a 12% difference in readmission between ADU and CRT service users. We found the readmission rate and the difference in admission rate were lower than we accounted for in the sample size calculation. The difference between models 2 and 3 led to attenuation of the hazard ratio more in keeping with the presence of confounding than the loss of power when adjusting for the additional characteristics in model 3. The differences in satisfaction, wellbeing and depression, each favouring ADUs, were all statistically significant with convincing effect sizes.

The fact that participants were not randomised could be seen as a limitation, however we were able to recruit a larger sample size than would have been likely in an RCT, and in the analysis we adjusted for a large number baseline differences between the two groups, including previous history of admission.

And fourthly, we collected a large quantity of data from routinely entered electronic health records, including our primary outcome, and as such were reliant on these records being accurate and up to date. We undertook a thorough cleaning process to correct obvious errors in this data, but this type of data is by nature not always very high quality.

### Implications for future research and practice

In future work it would be helpful to investigate why ADUs, despite being overwhelmingly popular with staff and service users, remain an underutilised model in the acute care pathway. Work to produce a model of best practice, along with service implementation guidelines, would provide a valuable resource to commissioners and service managers looking to increase choice for people in mental health crisis in their areas. Research about the place of ADUs in the complex mental health landscape would be beneficial, including how NHS services work with voluntary sector provision in this area.

It would also be helpful to generate further evidence regarding the factors that make these models so hard to implement in a sustained way. Making comparisons internationally may help to determine whether they are more sustained in less financially constrained service systems. Given the lack of recent RCT evidence about ADUs, and none at all comparing such units to other non-residential crisis services such as CRTs, a trial investigating different service models would be helpful to those planning, commissioning, and running acute care pathways.

## Conclusions

We have previously shown that the provision of ADUs in England is highly variable, with many parts of the country having no access to an ADU, and small numbers of service users benefiting from them (Lamb et al). However, where such services are available, service users and staff of ADUs value the units highly in terms of providing daily structure and contact with others, and emotional, practical and peer support [REFERENCE AD-CARE qual paper or NIHR report]. This cohort study provides evidence that people using ADUs had similar outcomes in terms of readmissions, and better outcomes for satisfaction, wellbeing, and depression, compared to CRT participants, after adjusting for baseline differences, and despite being a more severely unwell group. Those using ADUs were more likely to receive psychological input, a physical health check, and have carers involved, which are all interventions recommended by professional bodies and service user and carer groups.

## Data Availability

All data requests should be submitted to the corresponding author for consideration. Access to available anonymised data may be granted following review.

## Author details

Dr Danielle Lamb, Division of Psychiatry, UCL, UK (corresponding author)

Mr Thomas Steare, Division of Psychiatry, UCL, UK

Dr Louise Marston, Primary Care & Population Health, UCL, UK

Dr Alastair Cannaway, Clinical Trials Unit, University of Warwick, UK

Prof Sonia Johnson, Division of Psychiatry, UCL, UK, and Camden and Islington NHS Foundation Trust Dr James B. Kirkbride, Division of Psychiatry, UCL, UK

Dr Brynmor Lloyd-Evans, Division of Psychiatry, UCL, UK Dr Nicola Morant, Division of Psychiatry, UCL, UK

Dr Vanessa Pinfold, McPin Foundation, UK

Prof Scott Weich, School of Health and Related Research, University of Sheffield, UK

Prof David Osborn, Division of Psychiatry, UCL, UK, and Camden and Islington NHS Foundation Trust

## Required statements

### Declaration of Interest

None

### Funding

This work was funded by the National Institute for Health Research Health Services and Delivery Research Programme (project number 15/24/17). The views and opinions expressed therein are those of the authors and do not necessarily reflect those of the HS&DR Programme, NIHR, NHS or the Department of Health.

## Acknowledgements

We are very grateful to all the individuals who participated in our research: the staff, service users, and carers of the ADUs and CRTs in our study sites. We are particularly grateful to our PPI group, who provided valuable thoughts and comments on this work: Terry Harper, Hameed Khan, and Deb Smith. We are also grateful to the NHS R&D and NIHR CRNs (North Thames, West of England, Eastern, and West Midlands), especially the staff who facilitated and undertook data collection (special thanks to Lisa Carmody, Rose Tierney, Emily Benson, Anthony Wood, Tim Banks, Natalie Stonebank, Clare Davey, Ali Hodges, and Jo Morris).

## Author Contribution

DL ran the study, collected and analysed data, and wrote the first draft of this manuscript. TS collected data and provided comments on the manuscript. LM contributed to the data analysis and provided comments on the manuscript. AC, SJ, JBK, BL, NM, VP, and SW contributed to the conception and design of the study, and provided comments on the manuscript. DO conceived and designed the study, and provided comments on the manuscript.

## Notes

### Competing Interest Statement

The authors have declared no competing interest.

## References

1. Lloyd-Evans B, Osborn D, Marston L, Lamb D, Ambler G, Hunter R, et al. The CORE service improvement programme for mental health crisis resolution teams: results from a cluster-randomised trial. Br J Psychiatry. 2019;1–9.

2. Morant N, Lloyd-Evans B, Lamb D, Fullarton K, Brown E, Paterson B, et al. Crisis resolution and home treatment: stakeholders’ views on critical ingredients and implementation in England. BMC Psychiatry. 2017 Jul 17;17:254.

3. Weich S, Griffith L, Commander M, Bradby H, Sashidharan SP, Pemberton S, et al. Experiences of acute mental health care in an ethnically diverse inner city: qualitative interview study. Soc Psychiatry Psychiatr Epidemiol. 2012 Jan 1;47(1):119–28.

4. Marshall M, Crowther R, Sledge WH, Rathbone J, Soares-Weiser K. Day hospital versus admission for acute psychiatric disorders. Cochrane Database Syst Rev Online. 2011;12:CD004026.

5. Lamb D, Davidson M, Lloyd-Evans B, Johnson S, Heinkel S, Steare T, et al. Adult mental health provision in England: a national survey of acute day units. BMC Health Serv Res. 2019 Nov 21;19(1):866.

6. Attkisson CC, Zwick R. The client satisfaction questionnaire: Psychometric properties and correlations with service utilization and psychotherapy outcome. Eval Program Plann. 1982 Jan 1;5(3):233–7.

7. Haver A, Akerjordet K, Caputi P, Furunes T, Magee C. Measuring mental well-being: A validation of the Short Warwick–Edinburgh Mental Well-Being Scale in Norwegian and Swedish. Scand J Public Health. 2015 Nov 1;43(7):721–7.

8. Andresen EM, Malmgren JA, Carter WB, Patrick DL. Screening for Depression in Well Older Adults: Evaluation of a Short Form of the CES-D. Am J Prev Med. 1994 Mar 1;10(2):77–84.

9. Wing JK, Beevor AS, Curtis RH, Park SGB, Hadden J, Burns A. Health of the Nation Outcome Scales (HoNOS): Research and development. Br J Psychiatry. 1998 Jan;172(1):11–8.

10. Hardoon S, Hayes JF, Blackburn R, Petersen I, Walters K, Nazareth I, et al. Recording of Severe Mental Illness in United Kingdom Primary Care, 2000–2010. PLoS ONE [Internet]. 2013 Dec 12 [cited 2020 Aug 20];8(12). Available from: https://www.ncbi.nlm.nih.gov/pmc/articles/PMC3861391/

11. StataCorp. Stata Statistical Software: Release 16. College Station, TX: StataCorp LLC; 2019.

12. Werbeloff N, Chang C-K, Broadbent M, Hayes JF, Stewart R, Osborn DPJ. Admission to acute mental health services after contact with crisis resolution and home treatment teams: an investigation in two large mental health-care providers. Lancet Psychiatry. 2017 Jan 1;4(1):49–56.

13. Johnson S, Lamb D, Marston L, Osborn D, Mason O, Henderson C, et al. Peer-supported self-management for people discharged from a mental health crisis team: a randomised controlled trial. The Lancet. 2018 Aug 4;392(10145):409–18.

14. Priebe S, Jones G, Mccabe R, Briscoe J, Wright D, Sleed M, et al. Effectiveness and costs of acute day hospital treatment compared with conventional in-patient care - Randomised controlled trial. Br J Psychiatry. 2006;188:243–249.

15. Osborn DPJ, Lloyd-Evans B, Johnson S, Gilburt H, Byford S, Leese M, et al. Residential alternatives to acute in-patient care in England: satisfaction, ward atmosphere and service user experiences. Br J Psychiatry. 2010 Aug;197(S53):s41–5.

